# Evaluating AI-Assistance for Pathologists in Diagnosing and Grading Laryngeal Lesions

**DOI:** 10.1101/2023.07.23.23292962

**Authors:** Yaëlle Bellahsen-Harrar, Mélanie Lubrano, Charles Lépine, Aurélie Beaufrère, Claire Bocciarelli, Anaïs Brunet, Elise Decroix, Franck Neil El-Sissy, Bettina Fabiani, Aurélien Morini, Cyprien Tilmant, Thomas Walter, Cécile Badoual

## Abstract

**Importance:** Diagnosis of head and neck squamous dysplasias and carcinomas is challenging, with a moderate inter-rater agreement. Nowadays, new artificial intelligence (AI) models are developed to automatically detect and grade lesions, but their contribution to the performance of pathologists hasn’t been assessed.

**Objective:** To evaluate the contribution of our AI tool in assisting pathologists in diagnosing squamous dysplasia and carcinoma in the head and neck region.

**Design, Setting, and Participants:** We evaluated the effectiveness of our previously described AI model, which combines an automatic classification of laryngeal and pharyngeal squamous lesions with a confidence score, on a panel of eight pathologists coming from different backgrounds and with different levels of experience on a subset of 115 slides.

**Main Outcomes and Measures:** The main outcome was the inter-rater agreement, measured by the weighted linear kappa. Other outcomes on diagnostic efficiency were assessed using paired *t* tests.

**Results:** AI-Assistance significantly improved the inter-rater agreement (linear kappa 0.73, 95%CI [0.711-0.748] with assistance versus 0.675, 95%CI [0.579-0.765] without assistance, p < 0.001). The agreement was even better on high confidence predictions (mean linear kappa 0.809, 95%CI [0.784-0.834] for assisted review, versus 0.731, 95%CI [0.681-0.781] non-assisted, p = 0.018). These improvements were particularly strong for non-specialized and younger pathologists. Hence, the AI-Assistance enabled the panel to perform on par with the expert panel described in the literature.

**Conclusions and Relevance:** Our AI-Assistance is of great value for helping pathologists in the difficult task of diagnosing squamous dysplasias and carcinomas, improving for the first time the inter-rater agreement. It demonstrates the possibility of a truly Augmented Pathology in complex tasks such as the classification of head and neck squamous lesions.

## Introduction

Head and Neck Squamous cell carcinomas (HNSCC) are a significant global health concern, ranking sixth worldwide in both incidence and mortality rates ^1^. These cancers are notoriously associated with poor prognosis and high morbidity in the laryngeal and pharyngeal regions ^2,3^. One reason for these figures is the late diagnosis of invasive lesions and their dysplastic counterparts. Early detection of dysplasia is essential in preventing invasive carcinomas ^4^, and accurate grading is decisive as the grade remains the most important prognostic factor for the biological behavior of disease, guiding the physicians in their care strategy ^5^. Pathological examination is the gold standard diagnostic method ^6^ but poses many challenges. The small size of the samples impairs their optimal embedding orientation, often resulting in difficult-to-analyze tangent cuts. Changes in epithelium thickness between anatomical locations ^7^ and within a lesion itself ^8^ can make it challenging to differentiate reactive epithelial changes such as basal hyperplasia, from true dysplastic lesions. Moreover, dysplasia grading is a complex task that requires simultaneous consideration of multiple cytological and architectural features ^8,9^. Unlike most anatomical locations such as the uterine cervix or the digestive tract, the grading of head and neck dysplasia lacks immunohistochemical markers for guidance, thereby exclusively relying on Haematoxylin and Eosin (HE) staining and morphological assessment ^8,10,11^. The complexity of grading is evident from the multiple classifications proposed since the 1960s each with its own terminology (squamous intraepithelial neoplasia (SIN) by Friedmann and Osborn in 1976 ^12^, intraepithelial neoplasia of the larynx by Crissman and Fu in 1986 ^13^, laryngeal intraepithelial neoplasia (LIN) by Friedmann and Ferlito in 1988 ^14^, squamous intraepithelial lesion (SIL) by Gale *et al* in 2014 ^15^) and number of dysplasia categories ^11^. This multitude of options has led to ambiguity and dissonance without a single approach standing out as superior. Additionally, dysplastic lesions of the oral cavity are graded using a different system without a solid explanation for the rationale behind this distinction.

Numerous studies have highlighted the mediocre inter-rater agreement among pathologists, reflecting the significant challenges of pathological examination ^16,17^. In an attempt to address this issue, the WHO proposed in 2017 to simplify dysplasia grading by combining moderate and severe dysplastic lesions into a larger “high grade dysplasia” category ^10,15^. However, despite this simplification, reproducibility between pathologists remained unsatisfactory ^18^. Notably, the latest study on this topic by Mehlum *et al* ^19^ compared all reproducibility studies on head and neck squamous lesions in the literature, reported mediocre inter-rater agreement, demonstrating difficulties in providing reliable diagnosis even with the simplified binary system. Moreover, the scarcity of head and neck pathology specialists exacerbates the difficulties in getting optimal patient care. Therefore, the development of new tools to assist pathologists in their diagnoses is critical.

Recently, several studies have shown the benefits of Artificial Intelligence (AI) models for improved diagnostic accuracy and reproducibility among pathologists, leading to what could be called an “augmented pathology”. However, most of these works have focused on classification between different cancer subtypes or carcinoma gradings ^20–24^ . In a previous work ^25^, we proposed a deep learning model for classifying head and neck squamous lesions with an indication of the model’s confidence. However, we didn’t assess its effectiveness in a real-life setting.

The objective of the present study was to assess whether AI-Assistance could increase reproducibility among pathologists, ultimately leading to more effective and efficient management of patients, in the challenging task of detecting and grading laryngeal and pharyngeal (oropharynx and nasopharynx excluded) squamous dysplastic and invasive lesions.

## Materials and Methods

### Deep Learning Model

Based on the widely used Attention-MIL architecture ^26^, we developed, trained and validated a model for automatic grading of head and neck squamous lesions ^25^. For each slide, it generates two outputs: the predicted lesion (ranging from non-dysplastic, low grade dysplasia, high grade dysplasia, to carcinoma) and an associated confidence score. The confidence score is specifically designed to measure the model’s level of confidence for lesions on the same spectrum: it measures the extent to which the model hesitated with the second most probable (adjacent) class, as described in a previous work ^25^. The confidence threshold is optimized to reach an overall AUC > 0.9 on the validation set (thus settled at 0.5).

The model was trained using a dataset of 1949 digitized Haematoxylin, Eosin and Saffron-stained slides obtained from 456 patients who underwent either biopsies or surgical resection at Hôpital Européen Georges Pompidou (AP-HP, Paris, France). Each slide was associated with one class based on the most severe lesion present in the sample. The slides were digitized at 20X magnification using a Hamamatsu NanoZoomer® s360 scanner, resulting in a pixel resolution of 0.45 μm. To properly evaluate the model’s performance, an independent subsample of 115 biopsies was used as the test set. The classes for these slides were determined using a dual-blind review by two pathologists with expertise in head and neck squamous lesions, followed by a consensus meeting to thoroughly discuss any slides on which they disagreed. This reviewed portion of the dataset was used to evaluate the performance of the AI model Finally, the model was validated on an external dataset from another center (Hôpital Tenon, AP-HP, Paris, France) including 87 slides from 67 patients.

Details about the datasets are shown in **Supplementary Table A** and the performances of the standalone AI model are detailed in **Supplementary Table B**.

### Randomized Protocol and Pathologists Panel

The panel consisted of eight pathologists with varying experience levels and practice backgrounds: two residents in their last year of residency, three pathologists specialized in head and neck pathology, and three pathologists with no routine practice in head and neck pathology, as shown in **Figure 1A**. The two expert pathologists who labeled the 115 slides of the reference test set were not included in the panel. All panel members were tasked with reviewing the slides from the reference standard test set with and without AI-Assistance. Residents and non-specialized pathologists were provided with the references of the latest WHO grading system beforehand to update their knowledge. The study was designed as a randomized crossover trial, where each participant was randomly assigned to start with either the AI-assisted review or the non-assisted review (details in **Supplementary Figure A**), following protocols used in other studies ^22,27,28^. The pathologists independently reviewed the 115 slides and assigned a diagnosis to each of them without external input, in one uninterrupted session. A mandatory washout period of at least two weeks was required between the two reviews to avoid potential carryover effects.

**Figure 1.**
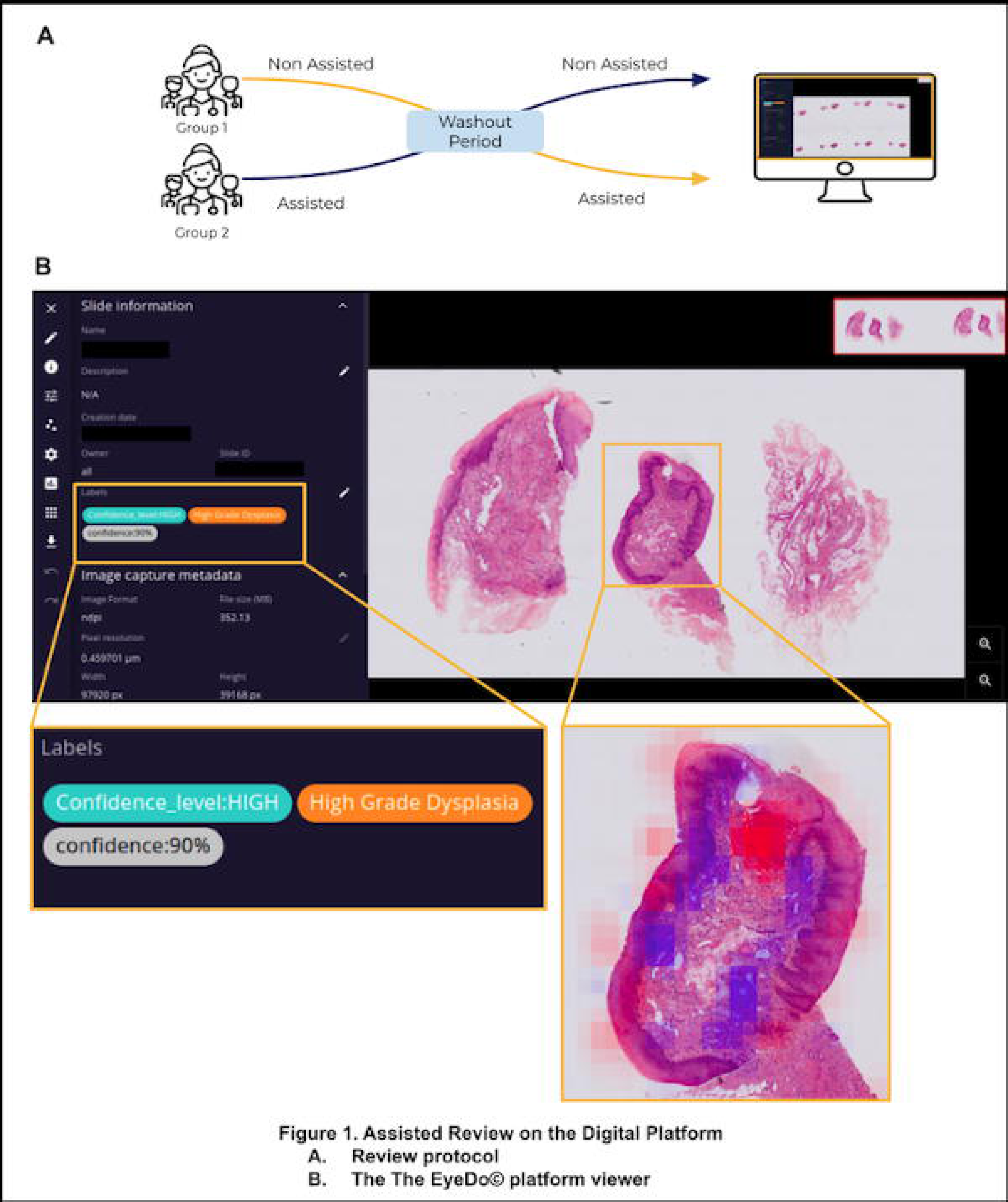
Assisted Review on the Digital Platform. A. Review Protocol The pathologists began either with assisted or non-assisted review, and switched after a washout period of at least two weeks. B. The EyeDo© platform viewer This platform provides to the pathologist, associated to the virtual slide, the model’s prediction, the confidence score expressed as a percentage, a categorization of the confidence (high or low) and a heatmap highlighting regions of the slide that contributed to the prediction

### Digital Platform and Review Process

The reviews were conducted using the EyeDo© digital platform (Tribun Health), a web-based viewer allowing for simultaneous visualization of the slides, the model’s prediction, and the confidence score, as shown in **Figure 1B**. A user guide was provided to the participants. Both the assisted and non-assisted reviews were performed on the same platform, but the slide names were changed between the two reviews to ensure blinding. During the non-assisted review, the pathologists had access only to the slides and were blind to any other information related to the case. For the assisted reviews, they were provided with the model’s prediction, the confidence score expressed as a percentage, a categorization of the confidence (high or low, following the threshold established beforehand) and a heatmap that could be toggled on and off, highlighting regions of the slide that contributed to the prediction, as shown in **Figure 1B** and **Supplementary Figure B**. For each slide, the reviewers were asked to fill in a table with their diagnosis, with the slide names pre-filled in the order of appearance on the platform.

### Statistical Analysis

After all the panel members completed the assisted and unassisted reviews, their diagnoses were compared to the reference internal test set. Cohen’s kappa with linear weights was used as the primary metric to measure the reproducibility, to compare it with the other studies published in the literature. Other standard classification metrics (accuracy, sensibility, specificity, negative and positive predictive values) were computed per class in a one-versus-rest manner. Confidence intervals of the AI algorithm model were computed using 10000 bootstraps. Statistical differences of the metrics between the AI-Assisted and the non-assisted reviews were assessed with a paired t-test. To account for possible bias in the reference standard of the internal test set, the pairwise agreement between all panel members individually were computed for the two reviews. All statistical analyses were performed using python (v3.6.9), pandas (v1.1.5), scikit learn (v1.2.0) and SciPy (v1.6.0).

## Results

### Agreement between the Pathologists with and without AI-Assistance

Agreement comparisons are presented in **Figure 2**. The results show that AI-Assistance significantly improved inter-rater agreement, as indicated by the reduced range of kappa values, as shown in **Figure 2A** (non-assisted review : linear kappa’s range from 0.576 to 0.742 ; assisted review : linear kappa’s range from 0.698 to 0.767). The mean linear kappa of the non-assisted review was similar to the standalone model (0.675, 95%CI [0.579-0.765]) whereas the assisted review outperformed the AI (mean linear kappa : 0.73, 95%CI [0.711-0.748], p < 0.001). When considering pairwise agreement within the panel without taking the reference standard labels into account, as shown in **Figure 2B**, the mean linear kappa was 0.616 (95%CI [0.597-0.637]) in the non-assisted review and 0.736 (95%CI [0.721-0.752]) in the assisted review (p < 0.001). These results show that AI-Assistance led to increased consistency in grading among the pathologists.

**Figure 2.**
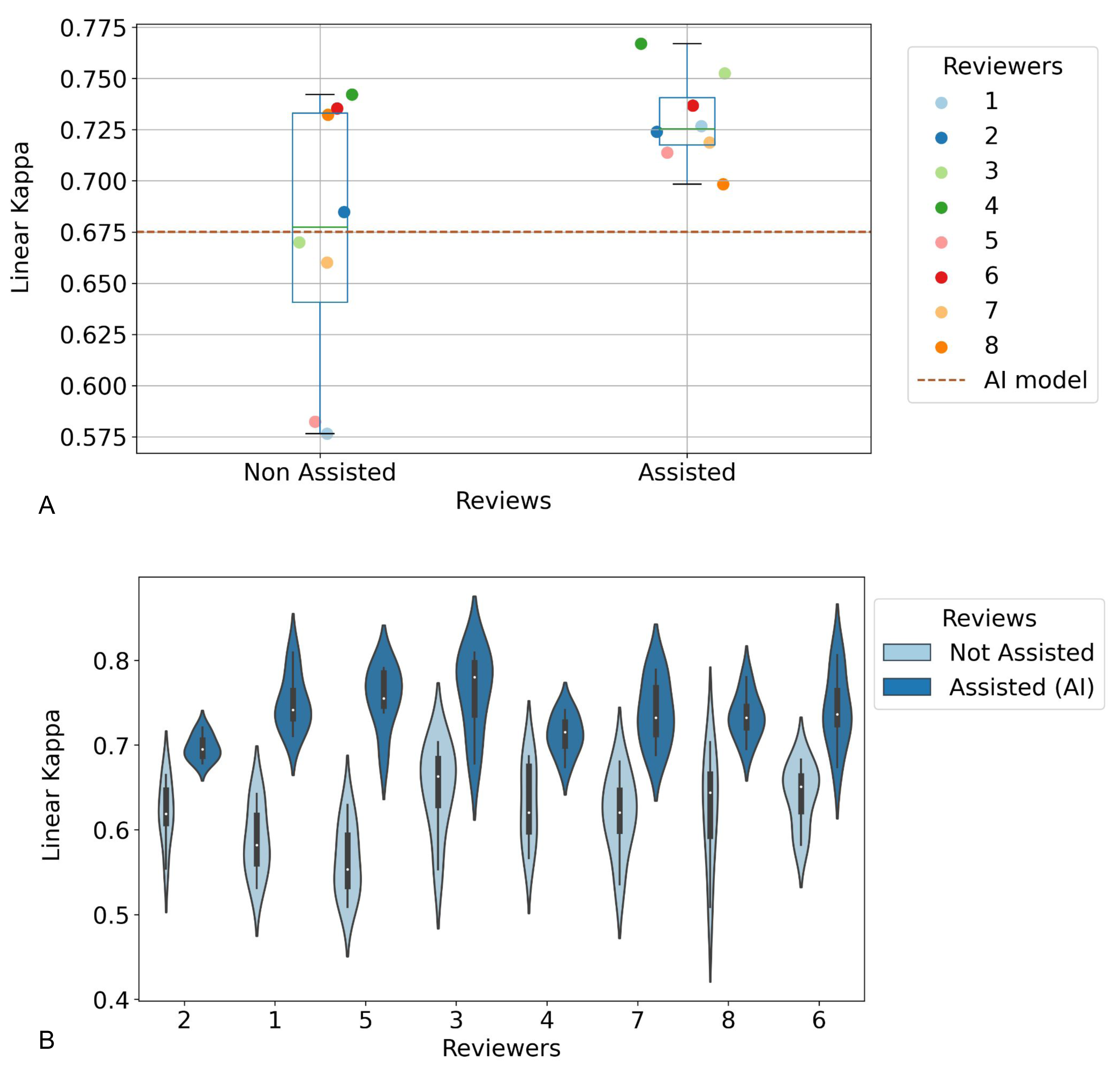
Agreement between Pathologists with and without AI-Assistance. A. Linear kappa values for each pathologist with and without the AI-Assistance, compared to the standalone AI-model. The assisted review significantly improved the inter-rater agreement and drastically reduced the kappa’s range between pathologists. B. Pairwise agreement showing an increase in the inter-rater agreement without considering the reference standard labels (linear kappa 0.734 assisted versus 0.619 non-assisted).

### Pathologists’ Performances Improvement with AI-Assistance

The overall performances of pathologists depending on their category (resident, non-HN specialist, HN specialist) are presented in **Figure 3**. The results demonstrate that the assisted residents and non-HN specialists outperformed the standalone AI model. Notably, their agreement became on par with those of HN specialists (linear kappas : residents with assistance : 0.725, 95%CI [0.723-0.728], non-HN specialists with assistance 0.744, 95%CI [0.713-0.776], HN specialists with assistance : 0.718, 95%CI [0.696-0.740]). Significant improvements were also obtained for the other metrics, as shown in **Supplementary Table C**. However, it is worth noting that the HN specialists, who already achieved better performances than the standalone model, did not benefit from much improvement from the AI-Assistance. These findings highlight the powerful impact of the AI-Assistance for non-HN specialists, especially valuable in the current situation of a lack of experts.

**Figure 3.**
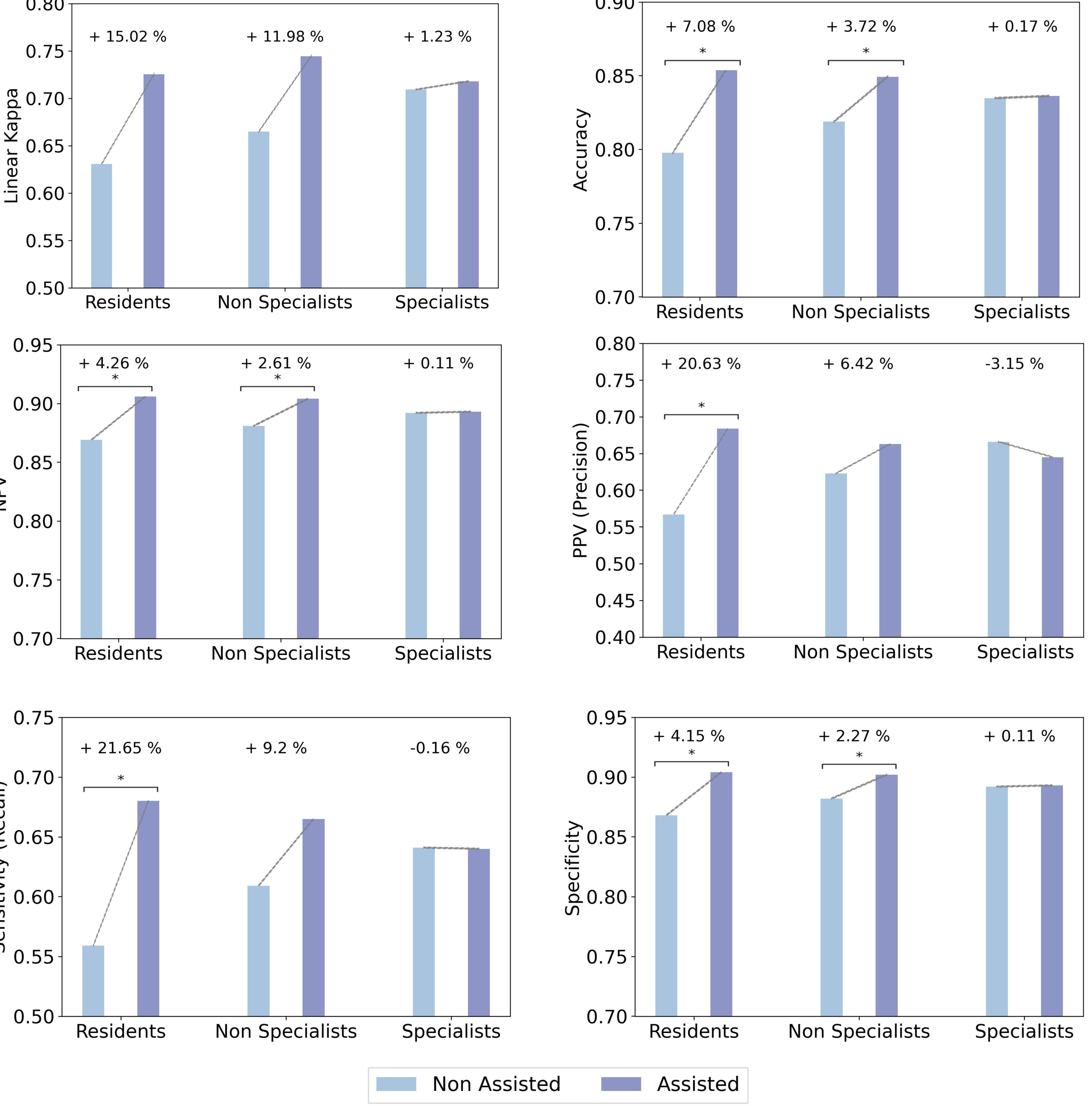
Performances Improvements with AI-Assistance per Pathologist Category. The assisted residents and non-HN specialists outperformed the standalone AI model and demonstrated significant improvement across all metrics. However, HN specialists didn’t benefit from much improvement.

### Impact of the Confidence Score on the Pathologists’ Performances

When considering the model’s confidence scores (as shown in **Figure 4)**, the results indicate that on high confidence predictions, reproducibility was significantly higher and with a reduced distribution of kappa values (high confidence predictions : linear kappa 0.809, 95%CI [0.784-0.834] for assisted review, versus 0.731, 95%CI [0.681-0.781] non-assisted, p = 0.018). There was no difference in the kappa values between assisted and non-assisted reviews when the confidence score was below the threshold (low confidence predictions: linear kappa 0.533, 95%CI [0.483-0.583] for assisted review, versus 0.522, 95%CI [0.459-0.586] non-assisted, p = 0.342), suggesting that the pathologists did not take the model’s predictions into account in these cases.

**Figure 4.**
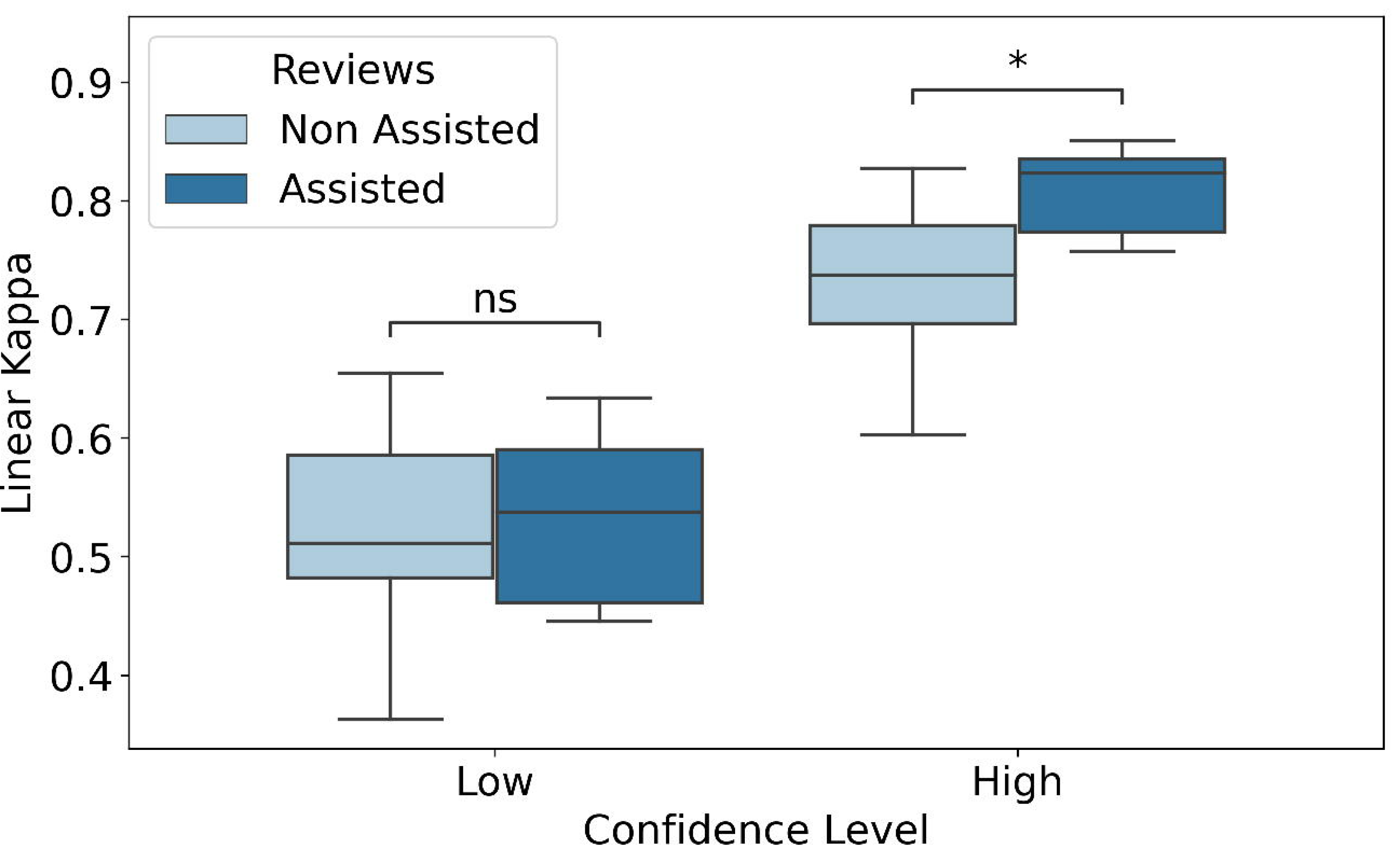
Pathologists’ Performances depending on the Confidence Level. The model’s confidence score guided the pathologists and improved the inter-rater agreement, with higher linear kappa on confident predictions (high confidence predictions: linear kappa 0.8 assisted versus 0.73 non-assisted, p < 0.001).

### Metrics Improvement with AI-Assistance depending on the Type of Lesion

Pathologists’ performances per diagnostic class are shown in **Table 1**. Globally, the pathologists were more performant with the AI-Assistance. Specificity improved significantly for low grade dysplasia, indicating better discrimination of this subtle lesion (non-assisted pathologists: 0.823, 95%CI [0.788-0.858] versus assisted pathologists: 0.867, 95%CI [0.854-0.880], p = 0.022). Moreover, performances showed drastic improvements for high grade dysplasia and carcinoma, the pathologists becoming more efficient than the standalone model. For instance, the accuracy for high grade dysplasia of the standalone model was 0.774, 95%CI [0.696-0.844] and 0.803, 95%CI [0.795-0.812] for the assisted review (p = 0.003). These results show that the AI tool assisted pathologists in identifying these harmful lesions with the highest therapeutic impact, and the combination of the pathologist’s expertise and the AI analysis proved to be complementary and more powerful when used together.

## Discussion

To the best of our knowledge, the benefit of an AI-Assistance in the difficult task of detecting and grading dysplasia in the laryngeal and pharyngeal regions wasn’t assessed before. Due to their limited number, studies on dysplasia detection and classification by AI models have not assessed whether these models enhance the performance of pathologists. Furthermore, almost all of them didn’t follow any official classification system. For instance, Tomita *et al* ^29^ decided to combine low grade and high grade dysplasia into a single class, and invasive adenocarcinoma and severe high grade dysplasia in another one, to address the lack of available data. Other works on Barrett’s esophagus ^30,31^ didn’t grade dysplasia either. Thus, a significant drawback of these studies is the limited size of their datasets, leading to insufficient training sets that struggle to identify subtle pathological features. In contrast, our model had the advantage of a comprehensive training set consisting of nearly 2000 slides and was further tested on two separate test sets. To our knowledge, only one previous work on gastric dysplasia successfully differentiated between epithelial regeneration change and dysplasia and graded the latter in a large cohort ^32^. Yet, this study didn’t evaluate if the AI tool could enhance the accuracy of pathologists in practical scenarios.

The deployment of an AI model, however powerful it may be, cannot fully replace the pathologist’s insight because of possible errors which could lead to harmful patient outcomes. Thus, pathologist validation of the AI predictions is mandatory. For the first time in the field of dysplasia grading, our findings demonstrate that pathologists supplemented with AI were more efficient than the standalone deep learning model. Notably, AI-Assistance significantly improved the accuracy and reproducibility of non-HN specialists and less experienced pathologists. Hence, AI-assistance enabled our heterogenous panel of pathologists to outperform the results from previous studies16 and to achieve performances fairly comparable to those of the panel of experts described in Gale *et al* ^15^, which achieved a weighted kappa of 0.80 among ten globally recognized expert pathologists. We show that in our case, the agreement was even greater on high confidence predictions, demonstrating that the confidence score is efficient in guiding non-specialized pathologists in their diagnosis. These results emphasize the need to integrate model confidence indicators in AI-assisted workflows.

The implications of our study are significant for clinical practice, since the use of our AI model by pathologists across varied backgrounds could lead to more precise and uniformed diagnoses, and improve patient care management. Additionally, the tool’s explanatory features, such as reliability scores and heatmap regions that influenced the prediction, provide the opportunity for pathologists to improve their own skills in the difficult task of dysplasia grading. This tool could be highly beneficial for the training of young pathologists, as well as for pathology departments lacking Head and Neck experts, especially in developing countries.

### Limitations

This study does have several limitations. Primarily, the model was trained using a single slide per sample, which was selected by a pathologist as the best representative of the lesion. In routine practice, a pathologist would examine multiple slides for the same sample, with a variation of the difficulty depending on the cut level. This selection of the best slide could have helped in the training of the model. However, all cuts on the selected slide were scanned and incorporated into the training and validation processes. Another limitation is the absence of clinical information, which is provided to the pathologist in a real-life setting. Finally, the relatively small size of the pathologist panel could have potentially restrained the statistical power of our tests. Despite these limitations, our results still highlight the great potential of AI-assisted pathology in the diagnosis of laryngeal and pharyngeal squamous lesions.

## Conclusions

In this study, we demonstrate the feasibility and effectiveness of authentic Augmented Pathology in the challenging task of diagnosing laryngeal and pharyngeal squamous lesions. For the first time, we describe a methodology that truly improves inter-rater agreement such as no classification system achieved before. By providing a reliable diagnosis and an efficient confidence score, we believe that our AI model has the potential for broad acceptance in clinical practice, thereby greatly improving patient care and management.

## FIGURE LEGENDS

**Supplementary Table B.** Standalone Standalone Model’s Performances. On the internal test set, the standalone AI model achieved an average AUC of 0.878 (95% CI: [0.801-0.937]) across the four classes, with an AUC > 0.9 for the detection of carcinoma, and an average linear kappa of 0.675 (95%CI: [0.575-0.760]). For the correct predictions, the confidence score had an average value of 0.846 +/-0.153, compared to 0.288 +/-0.150 for incorrect predictions, showing a good correlation between high model’s confidence and correct predictions. The overall AUC improved by 5.3% (0.931 [0.888-0.968]) when removing slides with low confidence. Conversely, overall AUC computed on the uncertain slides was 0.694 [0.580-0.797]. The linear kappa was 0.833 [0.737-0.905] on high confidence slides and 0.275 [0.077-0.449] on low confidence slides (+55.8%). Similar performances were observed on the external test set, where the model achieved an average AUC of 0.886 (95% CI: [0.813-0.947]).

|  |  | Diagnosis class |  |  |  |
| --- | --- | --- | --- | --- | --- |
| Metric | Status | Non-dysplastic | Low grade dysplasia | High grade dysplasia | Invasive carcinoma |
| Accuracy | Non assisted | 0.837 [0.813-0.861] | 0.751 [0.722-0.781] | 0.788 [0.765-0.811] | **0.902 [0.884-0.920]** |
|  | Assisted<br>(P-value*) | 0.866 [0.853-0.880]<br>(0.053) | 0.786 [0.770-0.802]<br>(0.068) | 0.803 [0.795-0.812]<br>(0.119) | **0.927 [0.911-0.943]**<br>(0.041) |
|  | Standalone model | 0.861 [0.791-0.922] | 0.757 [0.670-0.835] | **0.774 [0.696-0.844]** | **0.896 [0.835-0.948]** |
|  | Assisted<br>(P-value*) | 0.866 [0.853-0.880]<br>(0.224) | 0.786 [0.770-0.802]<br>(0.986) | **0.803 [0.795-0.812]**<br>(0.003) | **0.927 [0.911-0.943]**<br>(0.004) |
| Negative Predictive Value (NPV) | Non assisted | **0.882 [0.857-0.906]** | 0.864 [0.846-0.881] | 0.867 [0.843-0.891] | **0.916 [0.889-0.942]** |
|  | Assisted<br>(P-value*) | **0.917 [0.893-0.941]**<br>(0.022) | 0.869 [0.854-0.884]<br>(0.338) | 0.871 [0.849-0.893]<br>(0.384) | **0.944 [0.922-0.965]**<br>(0.029) |
|  | Standalone model | 0.911 [0.851-0.966] | 0.882 [0.802-0.95] | **0.819 [0.736-0.892]** | **0.921 [0.857-0.973]** |
|  | Assisted<br>(P-value*) | 0.917 [0.893-0.941]<br>(0.193) | 0.869 [0.854-0.884]<br>(0.942) | **0.871 [0.849-0.893]**<br>(0.000) | **0.944 [0.922-0.965]**<br>(0.048) |
| Positive Predictive Value (PPV)<br>(Precision) | Non assisted | 0.657 [0.580-0.734] | 0.382 [0.314-0.451] | 0.581 [0.534-0.627] | 0.880 [0.856-0.903] |
|  | Assisted<br>(P-value*) | 0.698 [0.660-0.736]<br>(0.206) | 0.438 [0.393-0.482]<br>(0.163) | 0.613 [0.591-0.634]<br>(0.075) | 0.899 [0.870-0.927]<br>(0.128) |
|  | Standalone model | 0.680 [0.480-0.864] | 0.400 [0.219-0.572] | **0.571 [0.333-0.800]** | **0.846 [0.722-0.946]** |
|  | Assisted<br>(P-value*) | 0.698 [0.660-0.736]<br>(0.406) | 0.438 [0.393-0.482]<br>(0.071) | **0.613 [0.591-0.634]**<br>(0.001) | **0.899 [0.870-0.927]**<br>(0.003) |
| Sensitivity (Recall) | Non assisted | **0.550 [0.440-0.660]** | 0.449 [0.367-0.530] | 0.608 [0.518-0.697] | **0.827 [0.765-0.889]** |
|  | Assisted<br>(P-value*) | **0.695 [0.600-0.790]**<br>(0.024) | 0.443 [0.370-0.516]<br>(0.546) | 0.612 [0.530-0.695]<br>(0.464) | **0.888 [0.842-0.933]**<br>(0.029) |
|  | Standalone model | 0.680 [0.517-0.862] | 0.545 [0.323-0.750] | 0.414 [0.240-0.593] | **0.846 [0.727-0.946]** |
|  | Assisted<br>(P-value*) | 0.695 [0.600-0.790]<br>(0.193) | 0.443 [0.370-0.516]<br>(0.527) | 0.612 [0.530-0.695]<br>(0.969) | **0.888 [0.842-0.933]**<br>(0.004) |
| Specificity | Non assisted | 0.917 [0.891-0.942] | **0.823 [0.788-0.858]** | 0.849 [0.815-0.883] | 0.941 [0.926-0.955] |
|  | Assisted<br>(P-value*) | 0.914 [0.892-0.936]<br>(0.583) | **0.867 [0.854-0.880]**<br>(0.022) | 0.868 [0.843-0.892]<br>(0.073) | 0.947 [0.930-0.965]<br>(0.306) |
|  | Standalone model | 0.911 [0.849-0.965] | 0.806 [0.725-0.882] | **0.895 [0.829-0.956]** | **0.921 [0.855-0.974]** |
|  | Assisted<br>(P-value*) | 0.914 [0.892-0.936]<br>(0.383) | 0.867 [0.854-0.880]<br>(0.071) | **0.868 [0.843-0.892]**<br>(0.000) | **0.947 [0.930-0.965]**<br>(0.011) |

\* one sided paired t tests. Results with statistical significance are in bold

## Ethics Approval and Consent to Participate

Our study was approved by the ethics committee of Assistance Publique - Hôpitaux de Paris Centre (CERAPHP. Centre - Institutional Review Board registration #00011928). All the patients were informed by a notification letter of the study and the possibility to refuse the use of their medical data, in line with current legislation. The study was performed in accordance with the Declaration of Helsinki.

## Data Availability Statement

The WSI dataset described in the manuscript were subject to hospital regulations and could not be publicly released. Data sharing with other research teams is possible under formal agreement with Assistance Publique - Hôpitaux de Paris (contact first and last authors for more information).

## Author contributions

Concept and design: CB, TW. Ethical approvals processes: CB, YBH. Creation of the clinical and pathology database, slides selection and labelling: YBH. Reference standard test set review: CB, YBH. Data management and processing, software implementation and statistical analyses: ML. Pathologists panel: CL, AB, CB, AB, ED, FEL, AM, CT. Results analysis: YBH and ML. Discussion of results: YBH, ML, CB, TW. Manuscript writing and review: all authors. All authors read and approved the final paper.

