## Supplementary Table A for "Evaluating AI-Assistance for Pathologists in Diagnosing and Grading Laryngeal Lesions"

| Characteristics | Training and Validation Sets | Reference Standard Test Set | External Set |
|---|---|---|---|
| **Number of patients** | 456 | 101 | 67 |
| Male | 376 (82.5%) | 79 (78.2%) | 14 (20.9%) |
| Female | 80 (17.5%) | 22 (21.8%) | 53 (79.1%) |
| **Number of samples** | 677 | 115 | 72 |
| Biopsies | 460 (67.9%) | 115 (100%) | 71 (98.6%) |
| Surgical resections | 217 (32.1%) | 0 (0%) | 1 (1.4%) |
| **Anatomical location** | | | |
| Larynx | 562 | 68 | 77 |
| Pharynx | 142 | 21 | 10 |
| **Number of slides** | 1949 | 115 | 87 |
| Biospies | 1370 (70.3%) | 115 (100%) | 86 (98.9%) |
| Surgical resections | 579 (29.7%) | 0 (0%) | 1 (1.1%) |
| **Diagnosis (worst lesions on the slide)** | | | |
| Non-dysplastic | 527 (27.0%) | 28 (24.3%) | 21 (24.1%) |
| Low grade dysplasia | 205 (10.5%) | 21 (18.3%) | 26 (29.9%) |
| High grade dysplasia | 602 (30.9%) | 30 (26.1%) | 17 (19.5%) |
| Carcinoma | 615 (31.6%) | 36 (31.3%) | 23 (26.4%) |
