## Supplementary Table B for "Evaluating AI-Assistance for Pathologists in Diagnosing and Grading Laryngeal Lesions"

| | | AUC [95% CI] | NPV [95% CI] | PPV (Precision) [95% CI] | Sensitivity (Recall) [95% CI] | Accuracy [95% CI] | Specificity [95% CI] | AUC (Precision/Recall) [95% CI] |
|---|---|---|---|---|---|---|---|---|
| **Reference Standard Set** | **Average** *(4 classes)* | **0.878 [0.801-0.937]** | **0.883 [0.811-0.945]** | **0.624 [0.439-0.795]** | **0.621 [0.452-0.788]** | **0.822 [0.748-0.887]** | **0.883 [0.815-0.944]** | **0.708 [0.543-0.844]** |
| | Non-dysplastic | 0.916 [0.854-0.962] | 0.911 [0.851-0.966] | 0.680 [0.480-0.864] | 0.680 [0.517-0.862] | 0.861 [0.791-0.922] | 0.911 [0.849-0.965] | 0.78 [0.612-0.903] |
| | Low grade dysplasia | 0.827 [0.735-0.898] | 0.882 [0.802-0.95] | 0.400 [0.219-0.572] | 0.545 [0.323-0.750] | 0.757 [0.670-0.835] | 0.806 [0.725-0.882] | 0.504 [0.29-0.706] |
| | High grade dysplasia | 0.827 [0.731-0.905] | 0.819 [0.736-0.892] | 0.571 [0.333-0.800] | 0.414 [0.240-0.593] | 0.774 [0.696-0.844] | 0.895 [0.829-0.956] | 0.633 [0.441-0.795] |
| | Carcinoma | 0.942 [0.884-0.982] | 0.921 [0.857-0.973] | 0.846 [0.722-0.946] | 0.846 [0.727-0.946] | 0.896 [0.835-0.948] | 0.921 [0.855-0.974] | 0.914 [0.827-0.973] |
| | All dysplasias | 0.869 [0.8-0.927] | 0.797 [0.697-0.889] | 0.745 [0.619-0.86] | 0.745 [0.615-0.863] | 0.774 [0.696-0.844] | 0.797 [0.692-0.887] | 0.849 [0.754-0.922] |
| **External Set** | **Average** *(4 classes)* | **0.886 [0.813-0.947]** | **0.883 [0.814-0.943]** | **0.655 [0.432-0.863]** | **0.629 [0.464-0.786]** | **0.822 [0.741-0.897]** | **0.878 [0.796-0.946]** | **0.722 [0.543-0.873]** |
| | Non-dysplastic | 0.89 [0.815-0.948] | 0.833 [0.747-0.916] | 0.6 [0.333-0.857] | 0.429 [0.217-0.647] | 0.793 [0.701-0.885] | 0.909 [0.836-0.971] | 0.65 [0.427-0.842] |
| | Low grade dysplasia | 0.799 [0.7-0.887] | 0.818 [0.705-0.909] | 0.5 [0.324-0.667] | 0.615 [0.407-0.792] | 0.701 [0.598-0.793] | 0.738 [0.618-0.843] | 0.58 [0.381-0.791] |
| | High grade dysplasia | 0.863 [0.755-0.951] | 0.882 [0.805-0.947] | 0.727 [0.444-1] | 0.471 [0.231-0.706] | 0.862 [0.793-0.931] | 0.957 [0.9-1] | 0.674 [0.423-0.859] |
| | Carcinoma | 0.994 [0.981-1] | 1 [1-1] | 0.793 [0.625-0.929] | 1 [1-1] | 0.931 [0.874-0.977] | 0.906 [0.83-0.971] | 0.983 [0.94-1] |
| | All dysplasias | 0.831 [0.74-0.908] | 0.727 [0.581-0.854] | 0.721 [0.587-0.848] | 0.721 [0.581-0.842] | 0.724 [0.621-0.816] | 0.727 [0.58-0.854] | 0.82 [0.702-0.911] |
