## Supplementary Table C for "Evaluating AI-Assistance for Pathologists in Diagnosing and Grading Laryngeal Lesions"

| Metric | Reviewers | Non Assisted [95% CI] | Assisted [95% CI] | P-value * |
|---|---|---|---|---|
| Linear kappa | Standalone AI | / | 0.675 [0.578-0.765] | / |
| | Residents | 0.631 [0.524-0.737] | 0.725 [0.723-0.728] | 0.168 |
| | Non-HN specialists | 0.665 [0.574-0.755] | 0.744 [0.713-0.776] | 0.061 |
| | HN specialists | 0.709 [0.661-0.758] | 0.718 [0.696-0.740] | 0.388 |
| Accuracy | Standalone AI | / | 0.822 [0.748-0.887] | / |
| | **Residents** | **0.798 [0.745-0.851]** | **0.854 [0.816-0.892]** | **0.004** |
| | **Non-HN specialists** | **0.819 [0.781-0.857]** | **0.849 [0.810-0.889]** | **0.015** |
| | HN specialists | 0.835 [0.801-0.868] | 0.836 [0.805-0.868] | 0.424 |
| NPV | Standalone AI | / | 0.883 [0.811-0.945] | / |
| | **Residents** | **0.869 [0.835-0.903]** | **0.906 [0.877-0.934]** | **0.013** |
| | **Non-HN specialists** | **0.881 [0.859-0.903]** | **0.904 [0.872-0.936]** | **0.048** |
| | HN specialists | 0.892 [0.875-0.908] | 0.893 [0.876-0.909] | 0.453 |
| PPV (Precision) | Standalone AI | / | 0.624 [0.439-0.795] | |
| | **Residents** | **0.567 [0.417-0.717]** | **0.684 [0.584-0.784]** | **0.011** |
| | Non-HN specialists | 0.623 [0.511-0.735] | 0.663 [0.550-0.776] | 0.08 |
| | HN specialists | 0.666 [0.558-0.774] | 0.645 [0.544-0.747] | 0.864 |
| Sensitivity (Recall) | Standalone AI | / | 0.621 [0.452-0.788] | / |
| | **Residents** | **0.559 [0.402-0.717]** | **0.68 [0.563-0.797]** | **0.034** |
| | Non-HN specialists | 0.609 [0.506-0.712] | 0.665 [0.531-0.8] | 0.108 |
| | HN specialists | 0.641 [0.547-0.734] | 0.64 [0.542-0.738] | 0.507 |
| Specificity | Standalone AI | / | 0.883 [0.815-0.944] | / |
| | **Residents** | **0.868 [0.826-0.911]** | **0.904 [0.880-0.928]** | **0.024** |
| | **Non-HN specialists** | **0.882 [0.845-0.918]** | **0.902 [0.874-0.93]** | **0.035** |
| | HN specialists | 0.892 [0.855-0.929] | 0.893 [0.866-0.919] | 0.479 |

* one sided paired t tests. Results with statistical significance are in bold
